# Validation of a Novel Smartphone Pupillometer Application Compared to a Dedicated Clinical Pupillometer in the Measurement of the Pupillary Light Reflex of Healthy Volunteers

**DOI:** 10.1101/2024.10.18.24315266

**Authors:** PM Middleton, W Davies, I Buzytsky

## Abstract

**Aim:** This study aimed to assess the correlation between the PLR curves of 25 healthy volunteers, generated with the MindMirror mobile telephone application, with PLR data from a dedicated clinical pupillometer.

**Materials & methods:** Paired pupillary light reflex curves were recorded from 25 healthy volunteers using the MindMirror mobile telephone application, and a Neuroptics NPi 200 clinical pupillometer. The curves were analysed for correlation using a Pearson correlation coefficient across both curve sets.

**Results:** Close correlation was demonstrated between all parameters except maximum and mean pupillary constriction velocity. These values were consistently lower in the MindMirror derived curves and may be explained by the intensity of the light stimulus available from a mobile telephone being significantly less that that used by the clinical pupillometer.

**Conclusion:** A mobile telephone equipped with a MindMirror mobile phone application may provide a reliable, low cost and widely available alternative to a clinical pupillometer in the assessment of the pupillary light reflex.

**Plain language summary:** Clinical pupillometry allows objective measurement of Pupillary Light Reflex (PLR) parameters, giving reproducible measurements shown to be independent predictors of adverse outcomes in patients with various neurological insults, and is increasingly being used as a clinical assessment tool among ambulatory patients outside of the hospital setting, particularly for concussion assessment in sports medicine. The MindMirror™ smartphone application uses artificial intelligence to identify and measure changes in important PLR components, and was shown to deliver variables with high levels of correlation and agreement with existing measurement tools such as the Neuroptics NPi-200TM Automated Pupillometer.

**Tweetable abstract:** A novel smartphone-based AI tool identifies potential predictors of neurological insults such as concussion with similar performance to dedicated tools.

## Introduction

The pupillary light reflex (PLR) is a response to changes in light levels where the pupil constricts or dilates. The size of the pupil is controlled by the iris sphincter muscle, which is innervated by the parasympathetic nervous system and causes the pupil to constrict, and the iris dilator muscle, which is innervated by the sympathetic nervous system and causes the pupil to dilate 1. This reflex is influenced by many factors and can be used to test for aspects of neurological function.

Pupil size and reactivity have been critical components of the clinical assessment of patients with alterations in level of consciousness, head injury and neurological disorders for decades. Pupillary examination may provide critical information related to new or worsening intracranial pathology, thereby facilitating prompt intervention and prevention of further neuronal damage.

Clinical pupillometry, which objectively measures pupillary parameters, has been shown to produce predictable and reproducible results across multiple measurements in response to changes in pathology and treatment. Abnormal values during a patient’s clinical course have been shown to be independent predictors of adverse neurological outcomes in patients with various neurological insults, including in cardiac arrest survivors, and in hemispheric cerebral infarction, subarachnoid haemorrhage, and traumatic brain injury. As a result, serial measurements via pupillometry have become a routine observation in neurocritical care units 2.

Automated clinical pupillometry, or Quantitative Pupillometry (QP), is increasingly being used as a clinical assessment tool among ambulatory patients outside of the hospital setting, particularly for concussion assessment in sports medicine. Pupillary responses have been shown to vary predictably in concussion patients, and as a sensitive biomarker of ongoing concussion effects may be useful in guiding return to play decisions. PLR metrics (maximum and minimum pupillary diameter, peak and average constriction / dilation velocity, percentage constriction, and time to 75% pupillary redilatation [T75]) were measured in 134 healthy control individuals and 98 athletes with concussion 3, at a median of 12 days following injury (interquartile range [IQR], 5 -18 days). The investigators found that eight of nine metrics were significantly higher among athletes with concussion, however there were sex-based differences observed as females with concussion exhibited longer T75 than males, and significantly diminished PLR metrics (e.g. smaller maximum pupil size) were observed after exercise.

There are several automated pupillometry devices available, including the Neuroptics NPi-200TM Automated Pupillometer. The Neuroptics NPi-200TM is a handheld device that measures pupil size and reactivity using infrared pupillometry, and generates a Neurological Pupillary Index (NPi) that incorporates multiple measurements into a single value for easy comparison and communication of results ^4^. Although existing infrared camera systems provide useful and precise measurements of the pupil, they are typically costly, use expensive disposable eyepieces and have limited availability.

Recently, mobile smartphone applications have gained attention in medicine for overcoming many of these limitations. The development of smartphone applications that can record and assess pupillary light reflex has been under way for some time, and has demonstrated the potential for accurate assessment of PLR ^5^. A smartphone-based infrared video pupillometer has been developed that compares favourably with commercially available devices, however this system requires external infrared light-emitting diodes and custom post-processing of test data ^6^.

Research has been conducted on artificial intelligence (AI)-based methods of accurately measuring the PLR and identifying changes in important components ^2^, however there is still disagreement about the validity and useability of many approaches. Before it is possible to deploy smartphone based, AI enabled technology to assess the pupillary light reflex of athletes suffering concussion, it is necessary to demonstrate that the technology can accurately record and plot pupillary light reflex curves. The current study addresses this by comparing the PLR curves of 25 healthy volunteers with PLR data from a NPi-200™ pupillometer.

## Materials and methods

### Study population

25 healthy adult volunteers were identified, and written consent was obtained. Subjects were asked to answer a brief questionnaire to identify any factors that may influence the PLR; these are recorded in table 1.

**Table 1.** Key PLR metrics as described in the NPi-200™ manual. MMCCP uses the same metrics and nomenclature to adhere to established standards.

| ID | Parameter | Description as used by NPi-200 |
| --- | --- | --- |
| 1 | Max Size | Maximum Diameter - Maximum pupil size before constriction |
| 2 | Min Size | Minimum Diameter - Pupil diameter at peak constriction |
| 3 | CV | Constriction Velocity - Average pupil diameter constriction velocity, measured in millimetres per second |
| 4 | MCV | Maximum Constriction Velocity - Maximum pupil diameter constriction velocity responding to the light stimulus, measured in millimetres per second |
| 5 | DV | Dilation Velocity - Average pupillary dilation velocity following the peak of constriction; the pupil recovers and dilates back to the initial resting size, measured in millimetres per second |

### Study protocol

The subjects were seated and had not undertaken any recent exertion. Matched PLR curves were obtained using a NPi-200™ pupillometer and the MindMirror™ application, which was installed on a Samsung Galaxy S21 FE™ smartphone. The PLR curves were recorded with both instruments as closely as possible in time. Data was uploaded to a secure cloud storage in the case of the MindMirror™ application. Data was downloaded from the eyepieces of the NPi-200™ using an Omnikey 5022™ and securely transmitted for comparison.

The MindMirror™ smartphone application uses the native camera and flash to record a video of the PLR, calculating a number of variables including average pupil diameter in pixels, maximum constriction and dilation velocity, and time in seconds for the pupil to reach 20%, 50% and 80% of the difference between baseline and minimum diameters. Figure 1.

**Figure 1.**
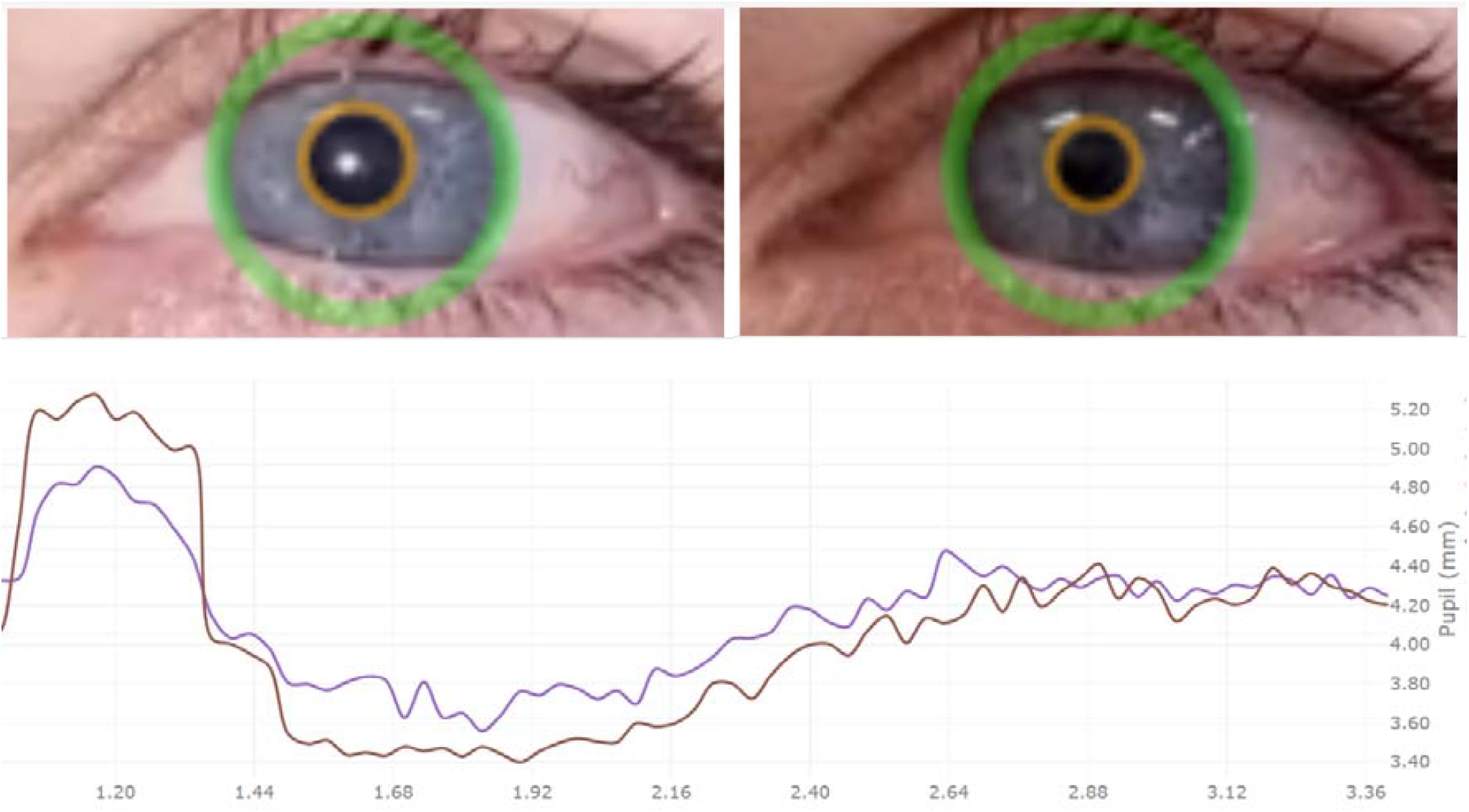
Image of MindMirror™ smartphone application’s capture of pupil constriction between flash at 1.2 seconds into the video, and achieving full constriction at 1.9 seconds into the video, or 0.7 seconds between flash and full constriction.

Using artificial intelligence (AI) algorithms, including machine learning and convolutional neural networks such as YOLO (You Only Look Once), specific to image recognition and classification and tasks that involve the processing of pixel data, the MindMirror™ application uploads the data to a highly secure cloud where processing takes place.

### NPi 200™ curve data extraction

Raw pupil diameter data from the NPi 200™ is currently inaccessible, therefore we prepared a NPi 200™ dataset by manually tracing key points over the screen capture of the device’s graphical output of pupil curves. We believe that this approach is methodologically sound since both the curve X/Y scales are referenced. Furthermore, the traced images were obtained using a very high resolution camera at 3000 by 4000 pixels, making the internal tracing graph scale as objectively accurate as possible. Figure 2.

**Figure 2.**
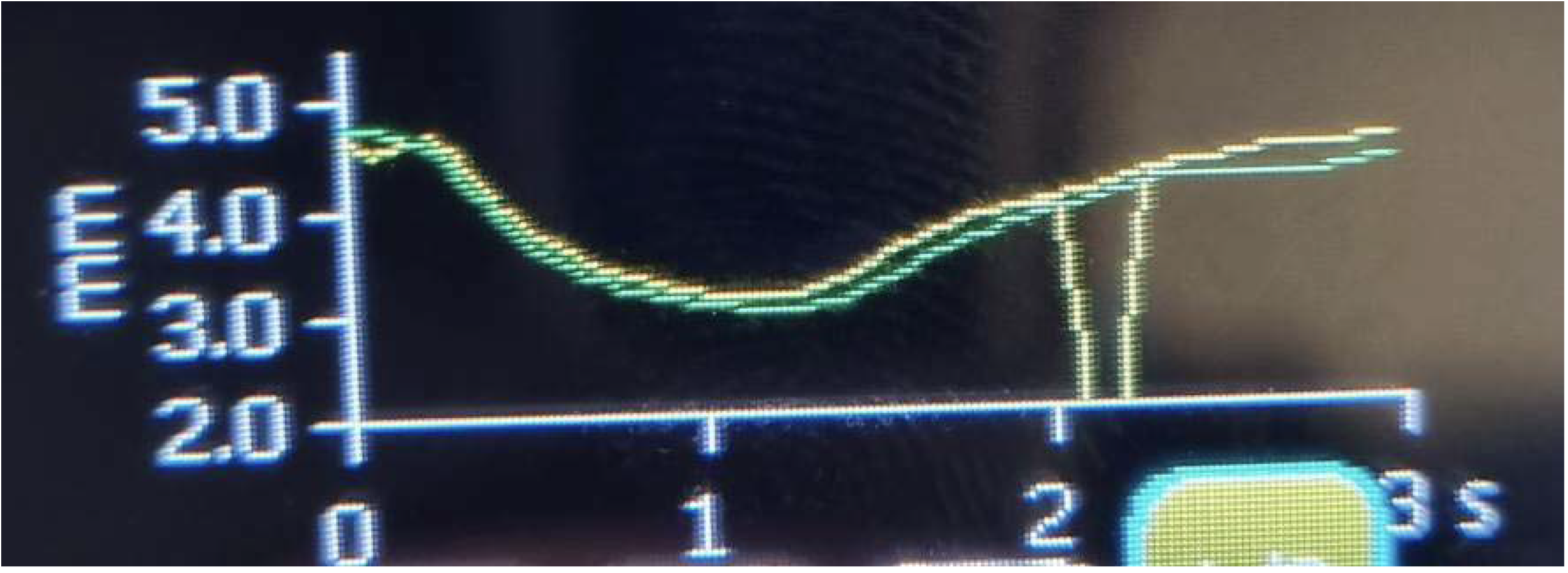
Screenshot of the NPi 200™ output produced at the end of the PLR measurement cycle of both eyes.

During tracing we chose to record key points at 30 frames per video, or 10 frames per second; this does not provide the level of detail compared to our captured rate of 30 frames per second with the MindMirror™ application, but gives an acceptable approximation of the curve shape and thus the key descriptive statistics.

### PLR Curve detection technology and workflows

The MindMirror™ application uses cloud computing to store, process, analyse and classify PLR curve information. The MindMirror™ Cloud Computing Platform (MMCCP) consists of a number of mutually complimentary technology solutions, or modules, each focusing on a particular aspect of PLR data analysis. The top-level design is represented in the diagram below, with descriptions of each technology module following. Figure 3.

**Figure 3.**
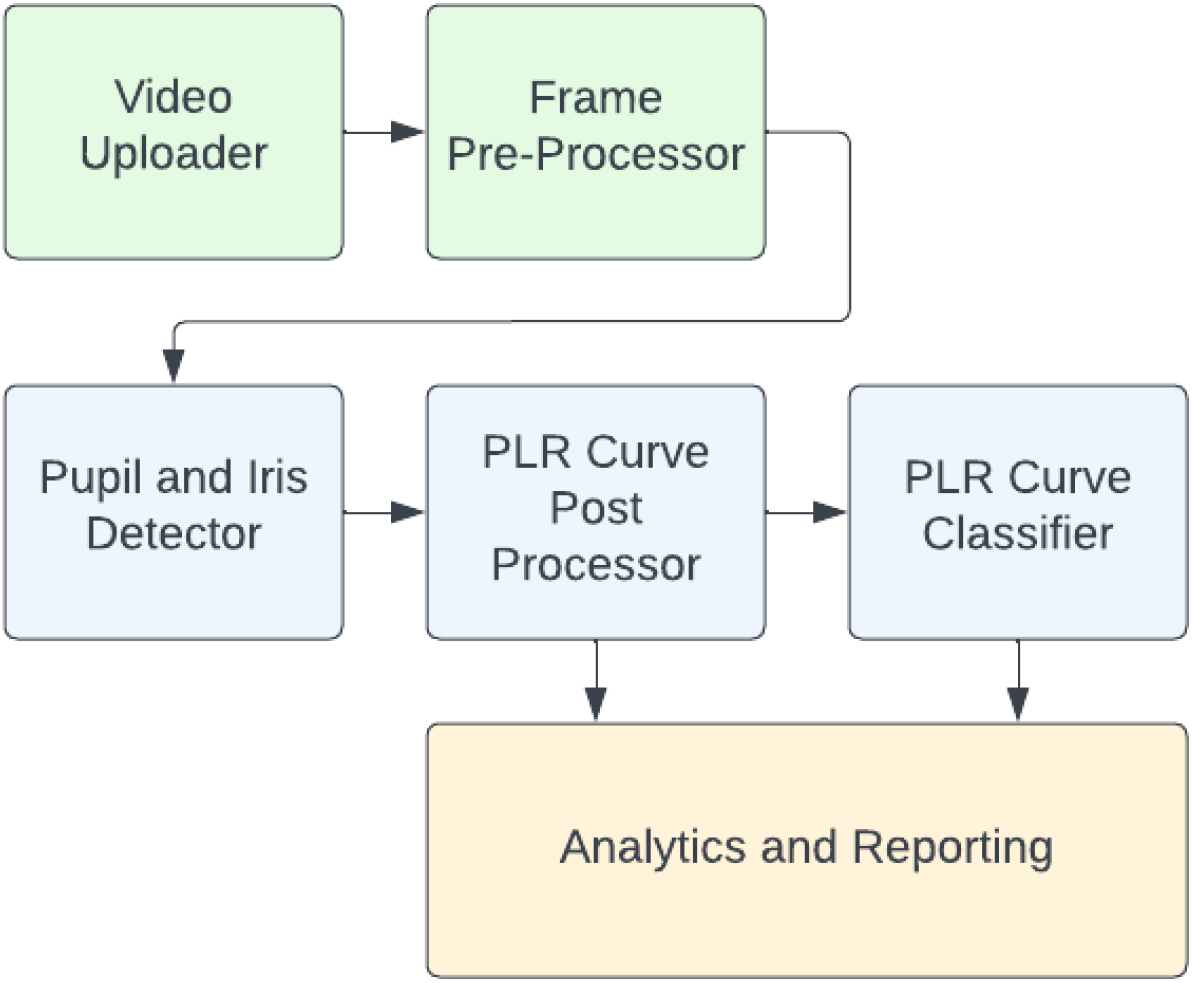
MMCCP key modules functional sequence diagram

### MMCCP Modules

1. Video Uploader
2. Pre-processor and Frame Extractor
3. Pupil and Iris Detector
4. PLR Curve Post Processor and Statistics Aggregator
5. Curve Classifier
6. Analytics and Reporting

### 1. Video Uploader

The Video Uploader module is responsible for secure upload of captured PLR videos and their associated metadata from the mobile device to the MMCCP. All data is encrypted in transport and is securely stored using Azure Cloud Storage to ensure regulatory compliance.

### 2. Pre-processor and Frame Extractor

Before PLR curve data can be detected and analysed, videos have to be scrubbed of any extraneous data, solely leaving test subject eyes as the focus of further analysis. In addition, isolating only portions of a test subject’s face where eyes are visible improves overall detection speeds and data volume requirements, and removes unnecessary personally identifiable information (PII). Figure 4.

**Figure 4.**
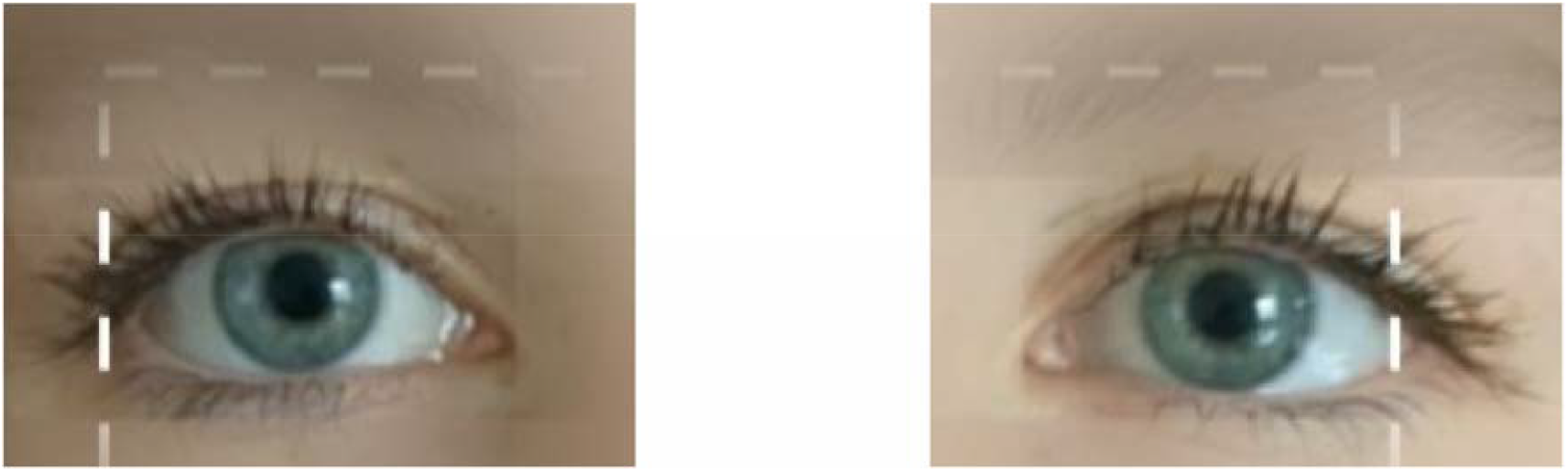
Example of a Point of Interest (POI) frame extraction.

### 3. Pupil and Iris Detector

The Pupil and Iris Detector is a key component of the MMCCP that uses machine learning models and object detection algorithms to identify the locations and sizes of test subject pupils and irises with a high degree of confidence. These algorithms are continuously revised using new video data to ensure that capture accuracy improves across multiple demographics, ethnicities, and lighting conditions. Figure 5.

**Figure 5.**
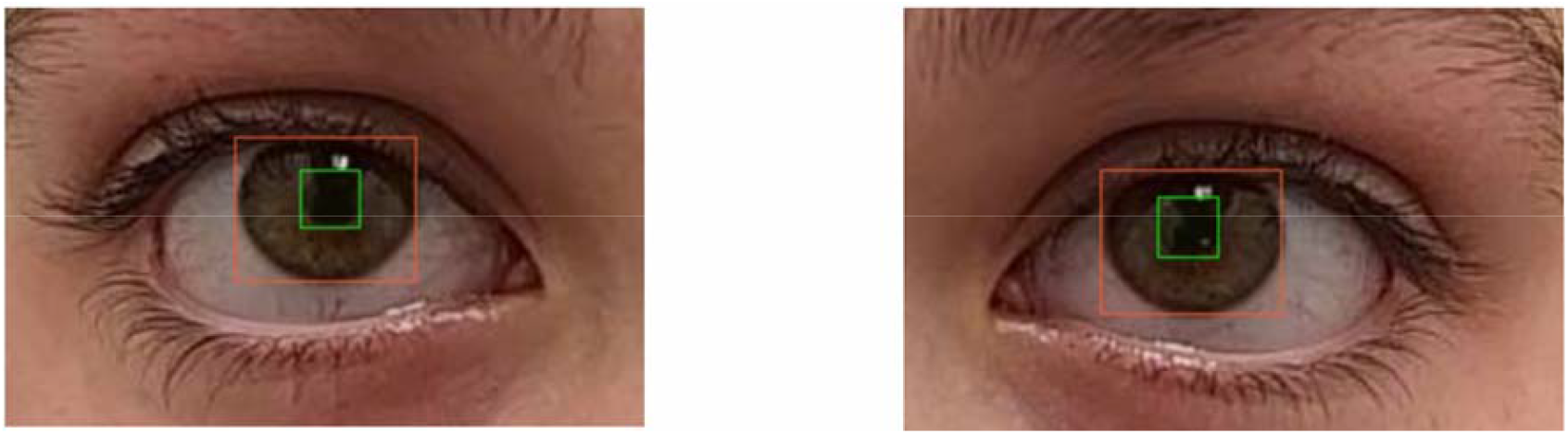
Pupil and Iris detection results via visualized bounding boxes

### 4. PLR Curve Post Processor and Aggregator

Pupil data is inherently noisy due to small object detection regression errors. To clean up some of this noise, an intelligent smoothing algorithm is applied to initial “raw” data to cleanly identify constriction and dilation statistics, when comparing such data to gold standard devices such as the NPi-200™. Figure 6.

**Figure 6.**
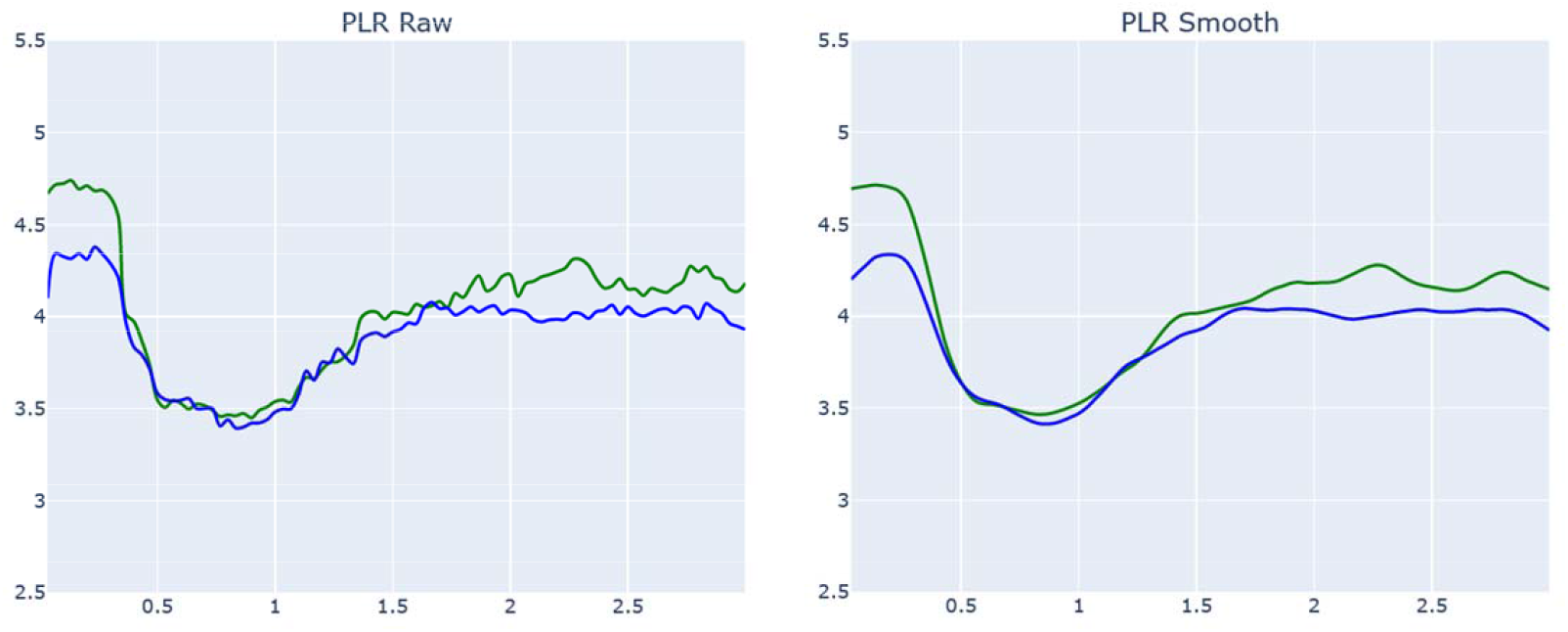
PLR curves in their original and smoothed representation

In this above example the raw PLR pupil diameter data is denoised / smoothed using an advanced mathematical algorithm. Once the pupil diameter data is appropriately de-noised, additional aggregate statistics are extracted from the curve time series, to allow for meaningful comparison of the MMCCP with the gold standard NPi-200™ aggregate outputs. The aggregate parameters that are computed by both MindMirror™ and NPi-200™ are summarised in Table 1.

In addition, the MMCCP computes and keeps track of parameters that are useful when building the PLR Curve Classifier models. Examples of such parameters are outlined in figure 7.

**Figure 7.**
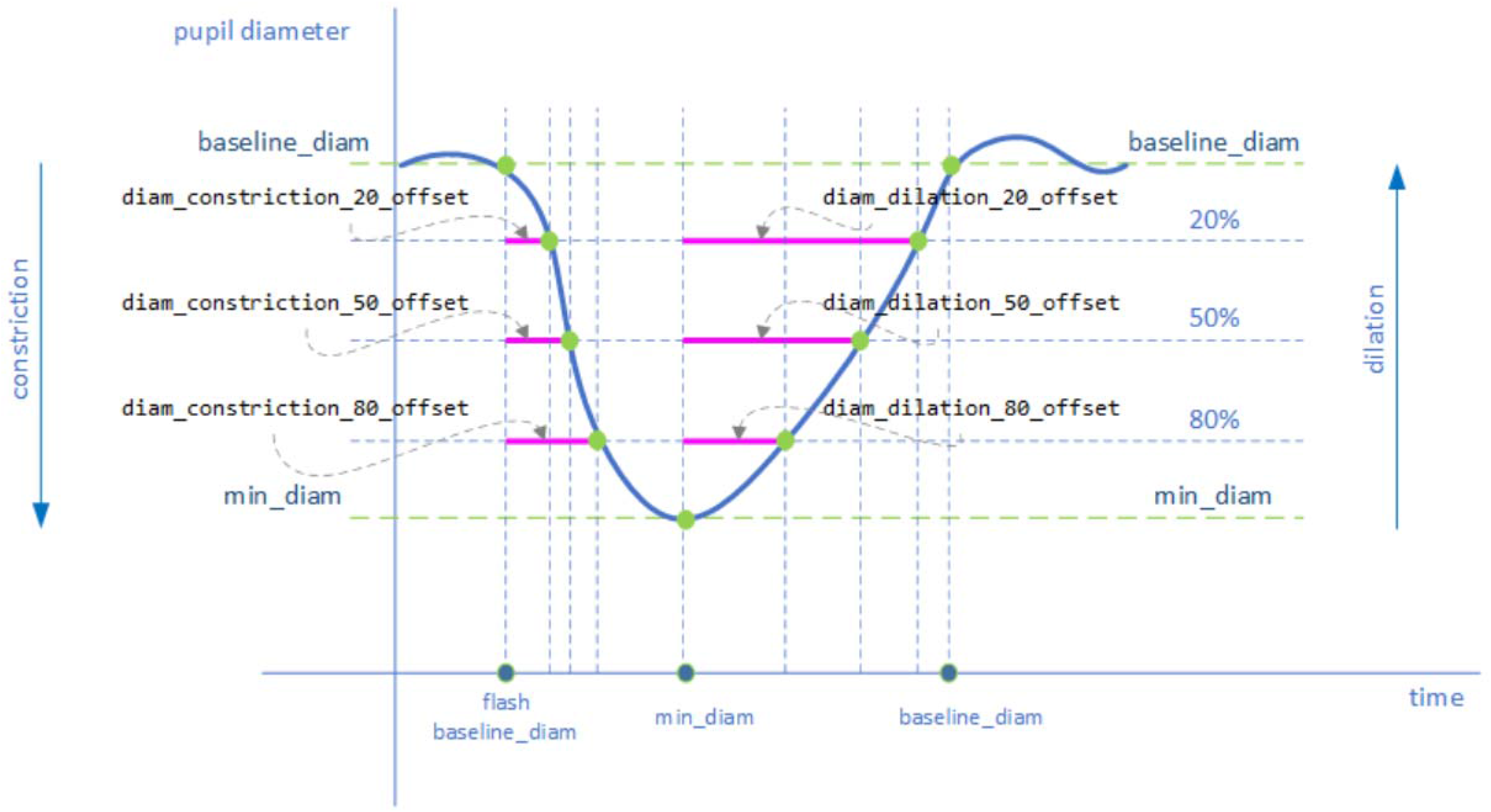
Additional PLR metrics computed by the MMCCP.

Finally, the MMCCP computes left / right pupil curve correlation scores, to demonstrate how closely left and right PLR curves resemble each other as demonstrated in figure 8.

**Figure 8.**
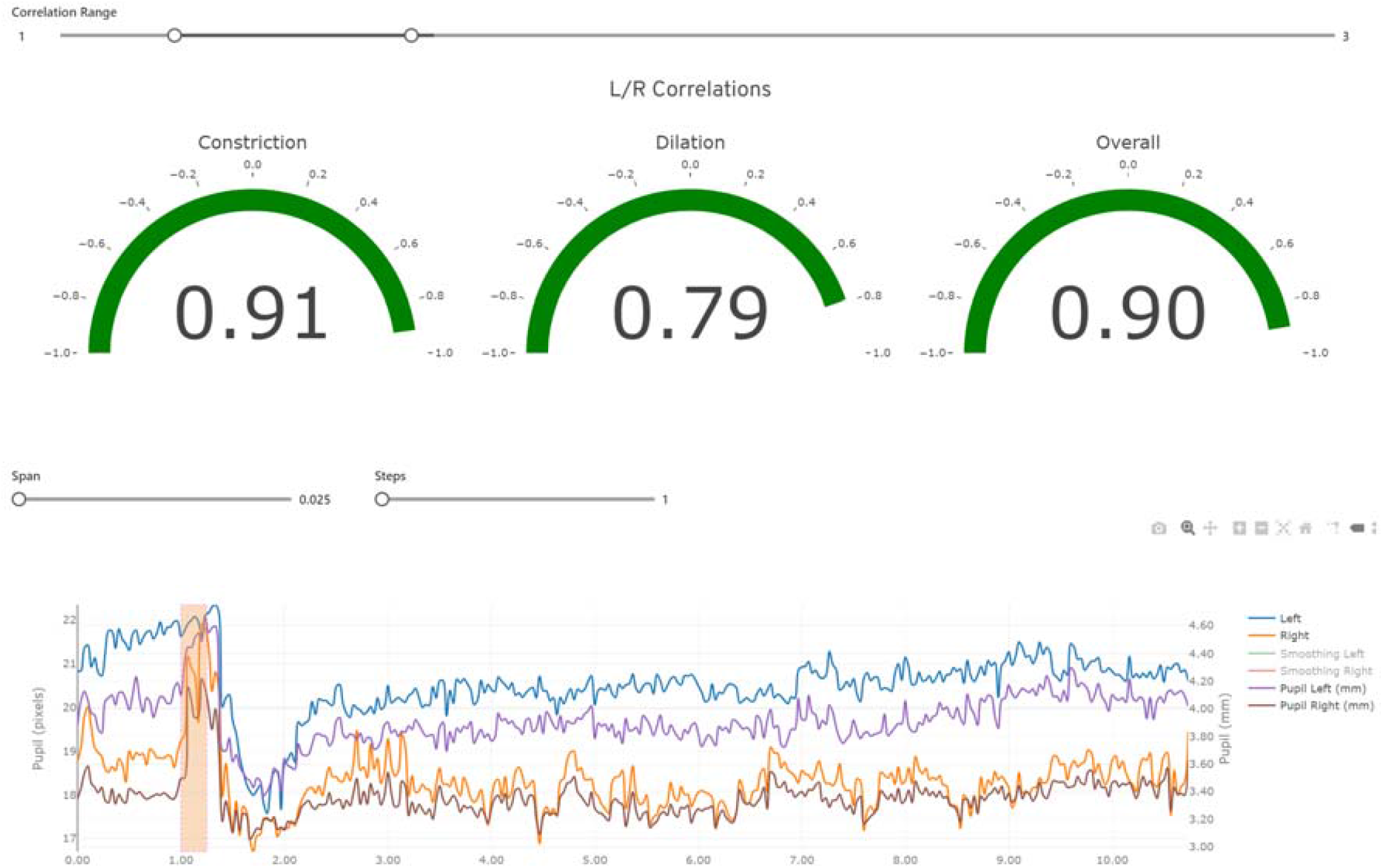
View of PLR chart and L/R correlation scores from the MindMirror Analytics Web Portal.

### 5. Curve Classifier

Once PLR curve data is collected and processed, additional analysis segmentation of the curves into “healthy” vs. “at risk” can be performed using another set of machine learning algorithms. By training different types of classifiers and using them in an ensemble formation, a high degree of confidence may be achieved when examining PLR curves that belong to a healthy cohort, compared to the curves of individuals with various at-risk conditions. Furthermore, the same approach may be utilised when tracking recovery after injury, especially with prior establishment of PLR baselines.

The Curve Classifier module provides additional datasets that may be associated with a test subject’s videos, and can be used for additional insights and research. The Classifier models are also continuously updated based on a stream of incoming PLR videos that have “healthy”, “at-risk”, or annotated curves demonstrating specific diseases such as concussion.

### 6. Reporting and Analytics

This module allows MMCCP users to utilise the collected PLR data to perform both visual and data analysis by reviewing curve shapes, key statistics, and the patient’s PLR history. In addition, these data may be shared with authorised third parties such as hospitals and medical professionals. An example can be seen in figure 9.

**Figure 9.**
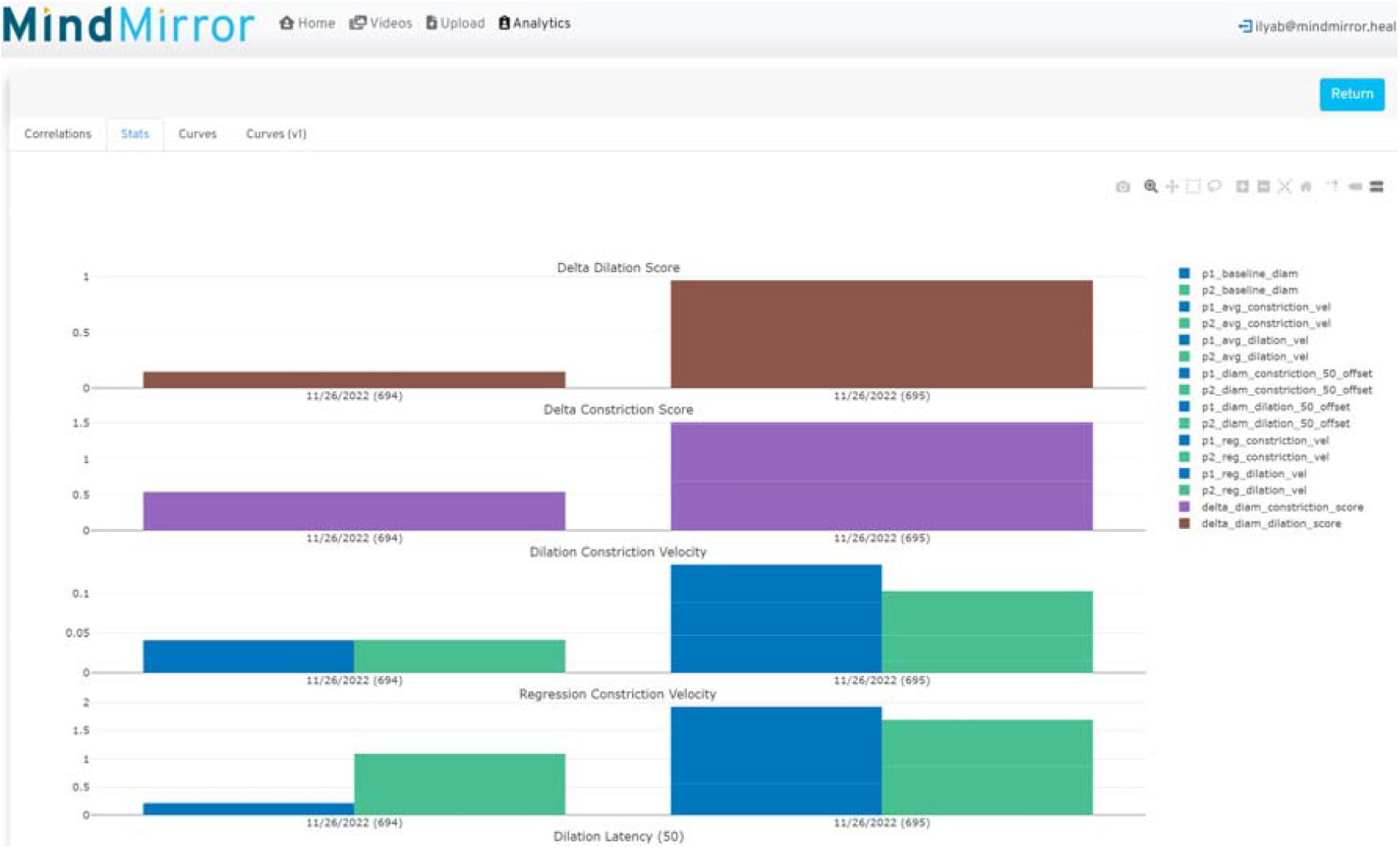
PLR statistics comparison page, available in the MindMirror Analytics Web Portal. For instance, by comparing scores over time, one can observe the evolution of a test subject’s recovery from injury.

## Results

### 1. Curve data preparation

To filter unavoidable noise resulting from pupil and iris bounding box coordinate detections, we applied smoothing algorithms to the pupil diameter time series data. In our case we used local regression, also known as LOESS (locally estimated scatterplot smoothing). Once the MindMirror™ curve data was smoothed, we aligned NPi 200™ and MindMirror™ PLR curves, ensuring that our time series represented the same events: the curves both commencing at the moment of flash, and analysed for the same 3 second interval following the flash. An example of comparative curves is shown on figure 10.

**Figure 10.**
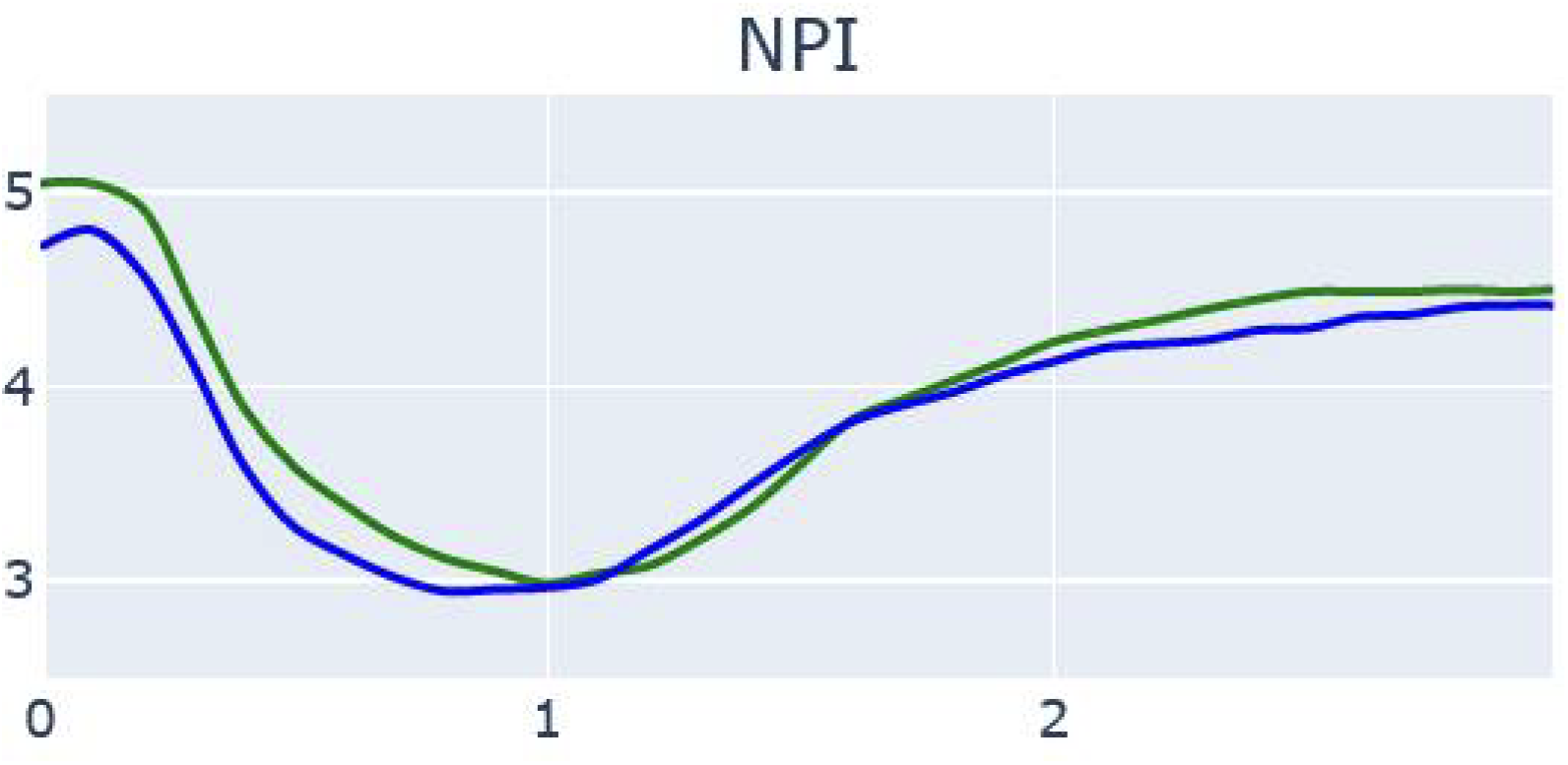
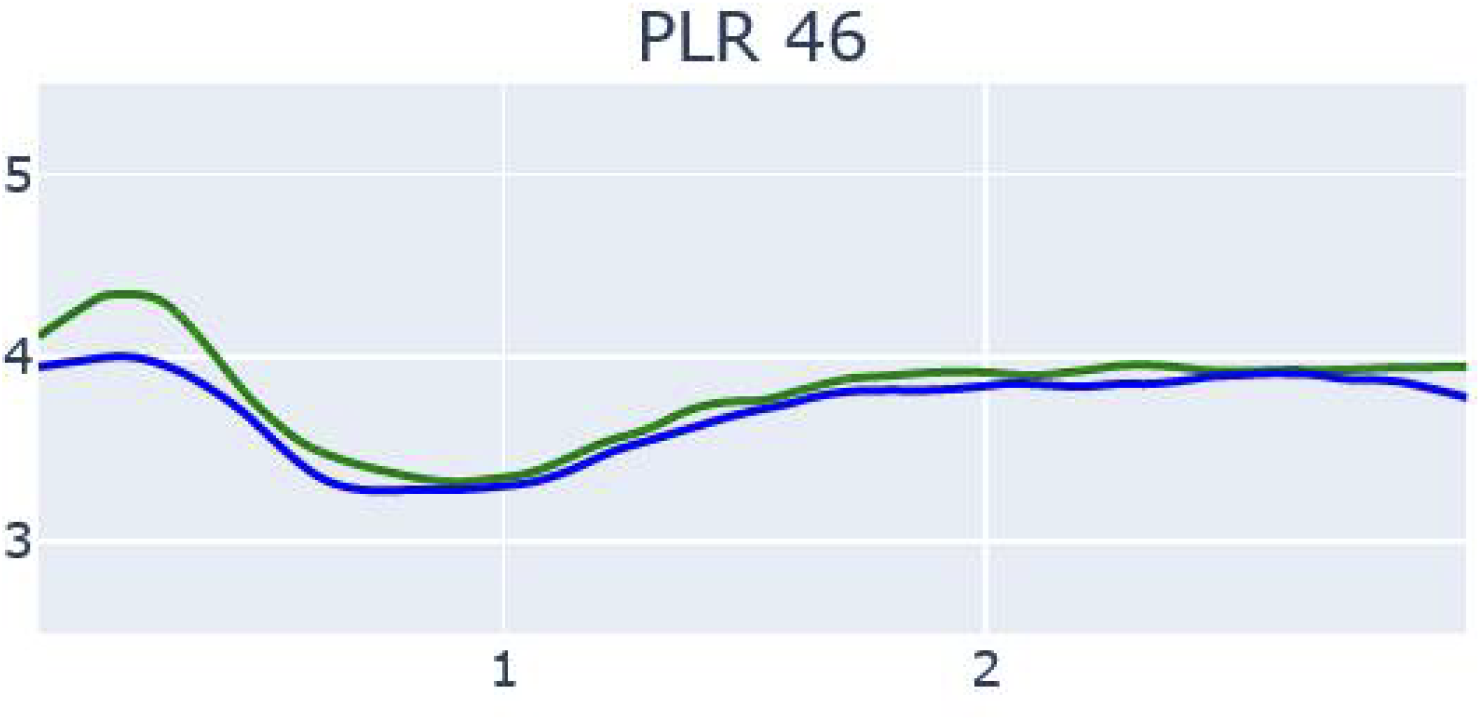
PLR curves of left and right eyes as captured by the NPi 200™ device (NPI) and MindMirror Mobile App, using a particular ML model to detect pupil and iris diameters (PLR 46)

### 2. Correlation analysis

To demonstrate similarity of the curve shapes across both methods of capture, we calculated a Pearson correlation coefficient across both curve sets. This showed a robust correlation across the entire curves; only two cases showed less than very good correlation, one of whom (vol 793) was identified as having a physical “blink” which interfered with measurement. Measured using the NPi 200™ and the MindMirror™ application in both right and left eyes, and during constriction and dilation, correlations coefficients were otherwise consistently above 0.8, with median values over 0.9 with narrow standard deviations. Individual test correlations are shown in Table 2 below.

**Table 2.** Pearson correlation coefficients between NPi 200™ curves and PLR curves detected by the MMCCP for the same patient taken sequentially in the same room.

| blob_id | study_id | L | R | LC | RC | LD | RD |
| --- | --- | --- | --- | --- | --- | --- | --- |
| 792 | C10004 | 0.495 | 0.476 | 0.714 | 0.721 | 0.915 | 0.987 |
| 793 | C10003 | 0.505 | 0.817 | 0.503 | 0.631 | 0.450 | 0.938 |
| 794 | C10001 | 0.748 | 0.826 | 0.883 | 0.845 | 0.972 | 0.985 |
| 795 | C10002 | 0.816 | 0.848 | 1.000 | 1.000 | 0.980 | 1.000 |
| 796 | C10006 | 0.877 | 0.853 | 0.976 | 0.952 | 0.983 | 0.955 |
| 797 | C10007 | 0.880 | 0.866 | 0.970 | 0.975 | 0.712 | 0.812 |
| 799 | C10005 | 0.886 | 0.886 | 0.961 | 0.911 | 0.953 | 0.982 |
| 801 | C10010 | 0.886 | 0.905 | 0.964 | 0.967 | 0.930 | 0.924 |
| 802 | C10011 | 0.904 | 0.908 | 0.980 | 0.975 | 0.945 | 0.915 |
| 805 | C10014 | 0.905 | 0.913 | 0.945 | 0.957 | 0.890 | 0.893 |
| 806 | C10015 | 0.912 | 0.923 | 0.964 | 0.938 | 0.982 | 0.941 |
| 807 | C10016 | 0.922 | 0.925 | 0.979 | 0.984 | 0.915 | 0.905 |
| 810 | C10018 | 0.932 | 0.936 | 0.990 | 0.975 | 0.980 | 0.976 |
| 815 | C10023 | 0.935 | 0.944 | 0.964 | 0.974 | 0.938 | 0.954 |
| 814 | C10022 | 0.937 | 0.945 | 0.934 | 0.957 | 0.957 | 0.962 |
| 813 | C10021 | 0.945 | 0.955 | 0.973 | 0.984 | 0.948 | 0.963 |
| 809 | C10019 | 0.978 | 0.956 | 0.974 | 0.974 | 0.972 | 0.983 |
| 808 | C10017 | 0.979 | 0.959 | 0.991 | 0.986 | 0.918 | 0.949 |
| 803 | C10012 | 0.982 | 0.963 | 0.985 | 0.981 | 0.907 | 0.977 |
| 798 | C10008 | 0.998 | 0.981 | 0.994 | 0.981 | 0.972 | 0.963 |
| Mean |  | 0.871 | 0.889 | 0.932 | 0.933 | 0.911 | 0.948 |
| Median |  | 0.909 | 0.918 | 0.971 | 0.974 | 0.947 | 0.958 |
| St Dev |  | 0.139 | 0.108 | 0.119 | 0.095 | 0.124 | 0.043 |

To illustrate concordance between methods at a granular level, we computed pairwise correlations between the NPi 200™ and the MindMirror™ application on a case-by-case basis in 20 subjects for which we collected hand traced NPI-200 data, analysing constriction and dilation velocities separately as seen above in Figure 11. Although discordance can be identified in a small number of pairs, 90% of correlations had values above 0.7, and more than 50% had values over 0.9, recognised as excellent correlation.

**Figure 11.**
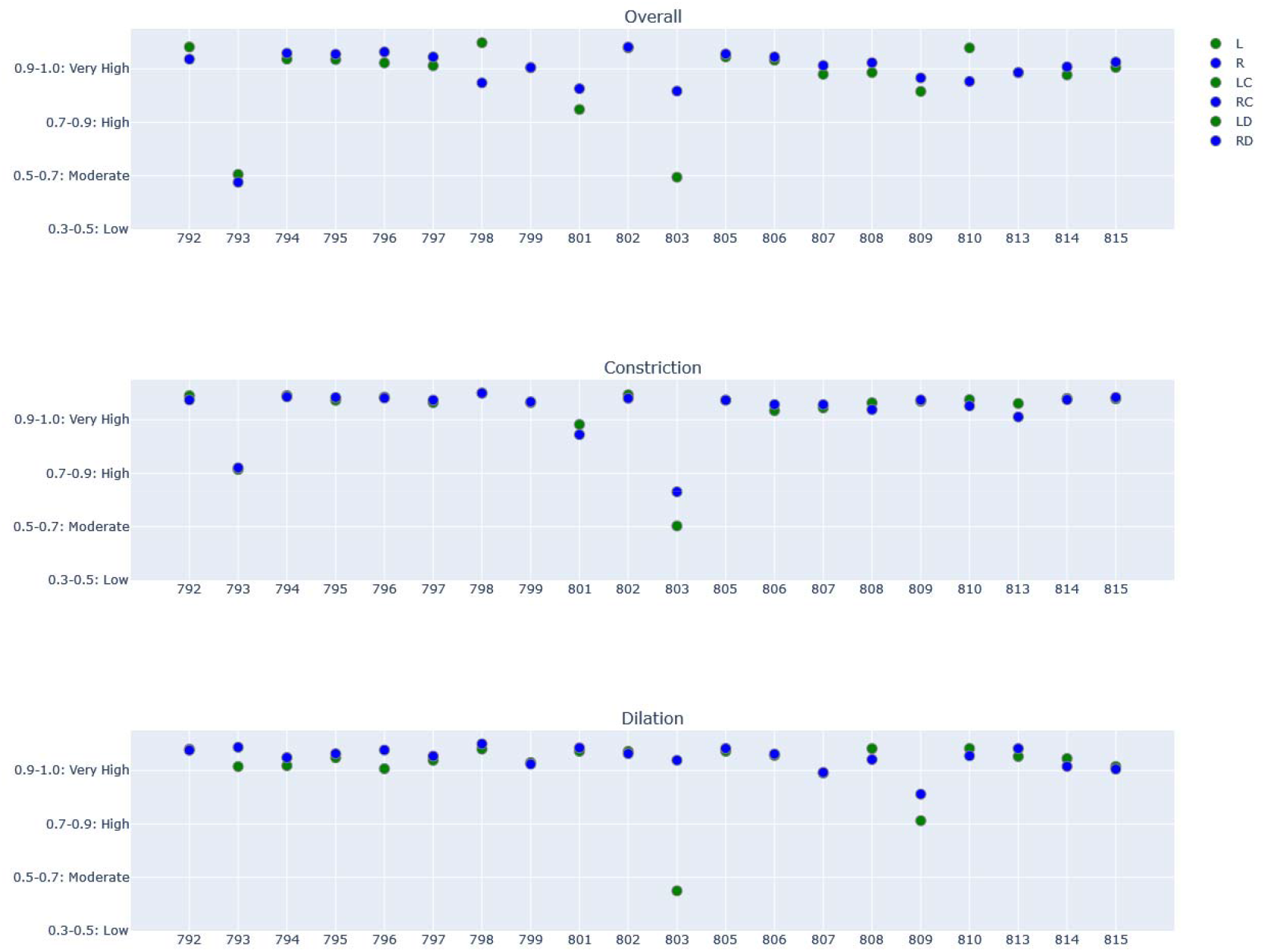
Full 3 second video, constriction and dilation Pearson correlation coefficients between NPi 200™ and MindMirror™.

### 3. Agreement analysis

We analysed agreement between NPi 200™ and MindMirror™ values for constriction velocity (CV), maximum constriction velocity (MCV) and dilation velocity (DV). Agreement is used to describe the differences between paired quantitative data, describing the same quantity but drawn from datasets derived from two separate methods of measurement. The intent is to ensure that a particular method used for quantitative measurement of a variable concerned is both reliable and reproducible for the intended use.

The mean difference between average pupil diameter Constriction Velocity measured by between NPi 200™ and MindMirror™ clustered around 0.5 mm s^-1^, with the MindMirror™ application consistently measuring velocity at a lower value. All but a single mean difference fell within the 1.96 SD limits of agreement. Maximum Constriction Velocity similarly demonstrated a mean difference between methods of approximately 1.0 mm s^-1^, with the MindMirror™ once again measuring the velocity at a lower value than the NPi 200™. Finally, the Dilation Velocity demonstrated very tight clustering around a mean difference of 0.2 mm s^-1^. This is demonstrated in figure 12. The individual parameters are shown in Table 3.

**Table 3.** Mean Constriction, Maximum Constriction and Mean Dilation velocities computed for NPI-200, MindMirror LOESS and MindMirror non-smoothed PLR curves.

|  | CV | CV | CV | CV | CV | CV | MCV | MCV | MCV | MCV | MCV | MCV | DV | DV | DV | DV | DV | DV |
| --- | --- | --- | --- | --- | --- | --- | --- | --- | --- | --- | --- | --- | --- | --- | --- | --- | --- | --- |
| type | mm | npi | mm_raw | mm | npi | mm_raw | mm | npi | mm_raw | mm | npi | mm_raw | mm | npi | mm_raw | mm | npi | mm_raw |
| eye | L | L | L | R | R | R | L | L | L | R | R | R | L | L | L | R | R | R |
| Video ID |  |  |  |  |  |  |  |  |  |  |  |  |  |  |  |  |  |  |
| 792 | 1.474 | 3.288 | 12.104 | 2.089 | 3.528 | 18.163 | 6.667 | 5.650 | 71.814 | 19.358 | 6.250 | 106.771 | 3.624 | 0.782 | 27.556 | 1.139 | 0.945 | 9.699 |
| 793 | 1.106 | 1.180 | 6.940 | 1.104 | 1.366 | 6.712 | 3.144 | 3.337 | 29.735 | 14.017 | 3.857 | 81.351 | 0.534 | 0.738 | 4.085 | 4.459 | 0.863 | 16.303 |
| 794 | 1.173 | 2.178 | 7.940 | 1.335 | 2.550 | 7.132 | 3.660 | 5.109 | 29.053 | 3.850 | 5.218 | 30.652 | 0.758 | 0.668 | 4.881 | 0.742 | 0.612 | 5.259 |
| 795 | 1.122 | 1.258 | 7.882 | 2.047 | 2.004 | 11.380 | 3.380 | 3.010 | 26.817 | 4.446 | 4.250 | 41.233 | 0.596 | 0.633 | 4.446 | 0.586 | 0.633 | 4.840 |
| 796 | 1.410 | 1.618 | 12.681 | 1.498 | 2.471 | 9.258 | 4.158 | 4.089 | 29.202 | 8.677 | 5.030 | 40.578 | 0.902 | 0.818 | 8.023 | 0.589 | 0.960 | 6.244 |
| 797 | 1.574 | 1.269 | 11.691 | 1.293 | 1.686 | 8.470 | 13.386 | 2.604 | 69.762 | 10.771 | 3.939 | 53.492 | 0.665 | 0.523 | 5.142 | 0.510 | 0.604 | 4.496 |
| 798 | 2.986 | 1.721 | 27.395 | 1.973 | 1.619 | 16.746 | 10.103 | 4.196 | 83.721 | 6.480 | 4.010 | 49.170 | 0.912 | 0.791 | 11.811 | 0.804 | 0.727 | 10.374 |
| 799 | 1.716 | 1.731 | 10.590 | 2.196 | 1.369 | 12.233 | 5.575 | 3.330 | 53.118 | 13.635 | 3.681 | 72.215 | 0.685 | 0.629 | 5.722 | 0.768 | 0.767 | 4.230 |
| 801 | 1.396 | 1.716 | 12.294 | 1.218 | 1.709 | 10.235 | 2.979 | 4.104 | 42.856 | 2.954 | 4.208 | 27.578 | 2.439 | 0.716 | 18.111 | 0.433 | 0.770 | 4.333 |
| 802 | 1.299 | 2.422 | 11.274 | 1.529 | 2.090 | 12.946 | 2.961 | 5.289 | 33.990 | 2.995 | 5.113 | 27.662 | 0.747 | 0.920 | 6.754 | 0.574 | 0.843 | 6.872 |
| 803 | 1.663 | 1.142 | 11.693 | 0.778 | 1.350 | 6.521 | 4.249 | 3.495 | 61.673 | 1.975 | 3.352 | 24.655 | 3.046 | 0.634 | 17.214 | 0.501 | 0.554 | 5.072 |
| 805 | 2.088 | 1.741 | 20.571 | 1.653 | 2.538 | 10.493 | 8.027 | 4.360 | 59.017 | 4.135 | 3.832 | 39.362 | 0.714 | 0.741 | 5.532 | 0.682 | 0.686 | 4.610 |
| 806 | 2.702 | 1.812 | 19.427 | 1.632 | 1.606 | 10.687 | 7.869 | 3.354 | 51.461 | 5.547 | 4.010 | 37.018 | 0.639 | 0.798 | 4.713 | 0.654 | 0.692 | 5.747 |
| 807 | 1.128 | 1.286 | 9.885 | 1.328 | 1.396 | 6.832 | 5.502 | 3.941 | 35.953 | 8.784 | 3.792 | 36.648 | 0.406 | 0.802 | 3.932 | 0.484 | 0.869 | 4.421 |
| 808 | 1.530 | 2.888 | 15.681 | 1.367 | 2.998 | 10.664 | 5.353 | 6.212 | 49.987 | 4.611 | 6.406 | 34.814 | 0.496 | 0.898 | 5.706 | 0.625 | 0.893 | 7.101 |
| 809 | 5.064 | 2.045 | 30.049 | 1.681 | 1.792 | 10.664 | 62.602 | 4.029 | 330.604 | 7.521 | 4.029 | 34.329 | 0.487 | 0.794 | 7.146 | 0.682 | 0.751 | 5.692 |
| 810 | 2.108 | 2.138 | 14.782 | 2.417 | 1.864 | 13.693 | 16.979 | 6.137 | 99.748 | 23.951 | 4.088 | 110.804 | 0.730 | 0.575 | 8.070 | 0.685 | 0.729 | 6.625 |
| 813 | 1.835 | 1.826 | 14.776 | 2.275 | 1.862 | 14.982 | 4.828 | 4.932 | 37.760 | 3.840 | 4.806 | 44.401 | 0.646 | 0.816 | 10.029 | 0.636 | 0.801 | 6.574 |
| 814 | 1.712 | 1.765 | 15.950 | 1.609 | 1.618 | 14.112 | 5.876 | 3.798 | 62.182 | 4.605 | 3.869 | 50.115 | 0.783 | 0.634 | 7.045 | 0.671 | 0.791 | 7.497 |
| 815 | 1.851 | 1.891 | 12.462 | 2.127 | 1.873 | 17.190 | 5.949 | 4.404 | 35.862 | 5.903 | 5.270 | 55.450 | 0.637 | 0.774 | 5.666 | 0.529 | 0.813 | 5.274 |
| Mean | 1.847 | 1.846 | 14.303 | 1.657 | 1.964 | 11.456 | 9.162 | 4.269 | 64.716 | 7.903 | 4.450 | 49.915 | 1.022 | 0.734 | 8.579 | 0.838 | 0.765 | 6.563 |
| SD | 0.907 | 0.552 | 6.056 | 0.439 | 0.582 | 3.540 | 13.081 | 1.010 | 65.670 | 5.826 | 0.846 | 24.699 | 0.898 | 0.105 | 5.982 | 0.866 | 0.113 | 2.844 |

**Figure 12.**
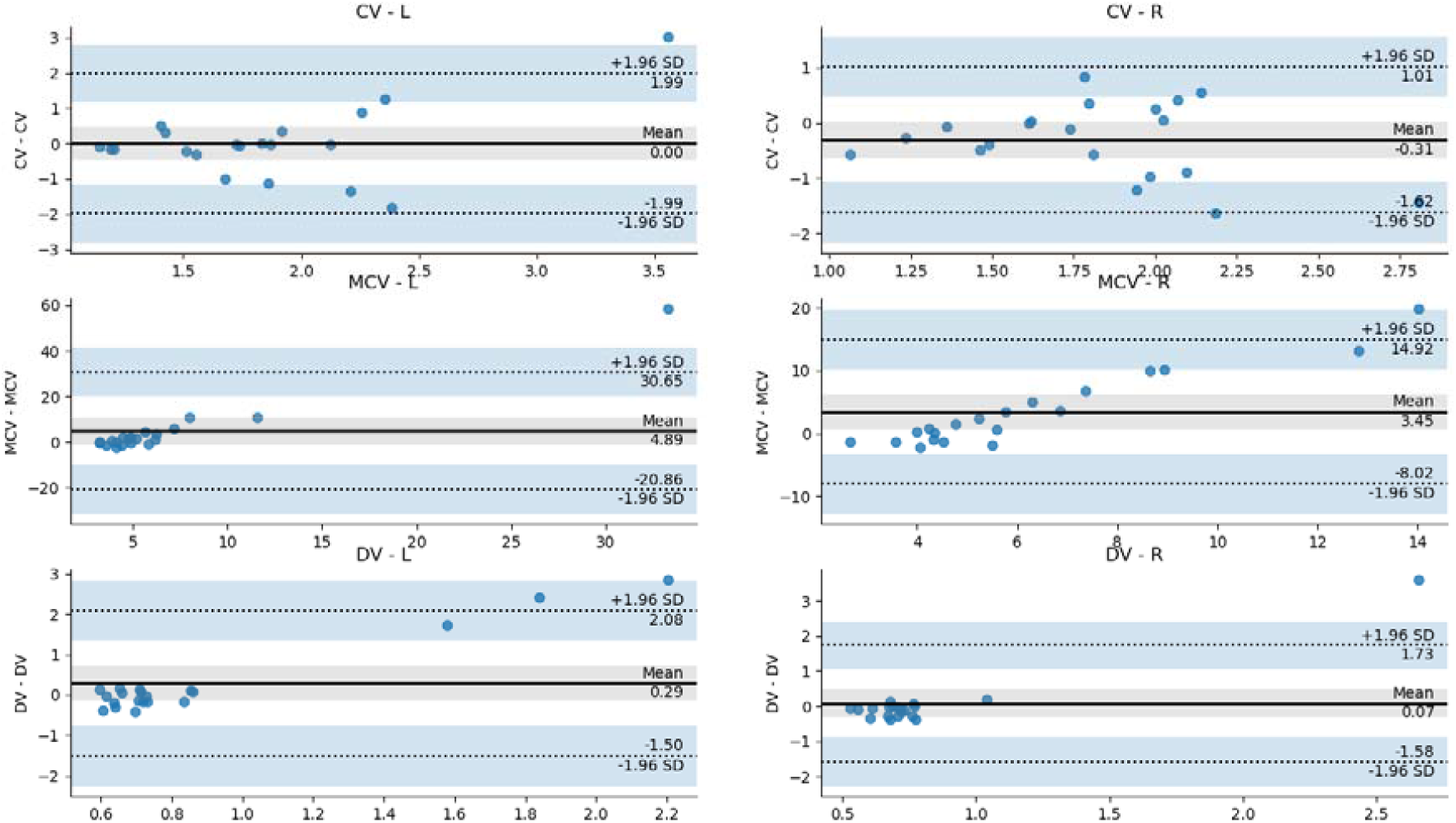
Bland–Altman plot (difference plot) showing agreement between NPi 200™ and MindMirror™ in constriction velocity (CV), maximum constriction velocity (MCV) and dilation velocity (DV).

## Discussion

Pupils exhibit distinct reactions to three different types of stimuli. They undergo constriction when exposed to bright light (the pupil light response), contract when focusing on nearby objects (the pupil near response) and dilate in response to heightened arousal and mental effort. Pupillary responses are partially reflexive, with a similar stimulus eliciting the same response, such as the pupils always constricting rather than dilating in reaction to light. Pupil responses also possess a voluntary component, influenced by higher-level cognitive processes.

The PLR is a fundamental aspect of level of consciousness assessment in multiple circumstances, particularly in traumatic brain injury, with abnormalities of pupillary response, or anisocoria, associated with neurological deterioration and secondary brain injury, and also correlated with poor neurological outcomes ^7 8^. Although pupil abnormalities have been demonstrated to be independent predictors of adverse neurological outcomes among patients with a variety of neurological insults, including cardiac arrest, hemispheric cerebral infarction, subarachnoid haemorrhage and traumatic brain injury, pupillary reflexes are also impacted by many potential confounding variables, such as ageing, sex, eye symmetry and smoking ^9^, Parkinson’s disease, Alzheimer’s disease, multiple system atrophy, schizophrenia, migraine, cluster headache, arthritis, Meniere’s disease, multiple sclerosis, heart failure, familial dysautonomia and generalised autonomic neuropathy ^10^. Increased parasympathetic nervous system activity associated with physical fitness has also been shown to affect multiple PLR parameters compared to controls ^11^, and PLR has been demonstrated to be affected by diabetes, ingested methadone, and other opiates ^12^, ACE inhibitors, and tricyclic antidepressants ^13^.

“Corectopia” or ectopia pupillae, was first discussed in 1907 by Wilson, reporting on cases where intracranial mass lesions appeared to impact on the autonomic nervous supply to the iris ^14^. However, in their 1974 paper describing the Glasgow Coma Scale, Teasdale and Jennett ^15^ observed “unstructured observations commonly result in ambiguities and misunderstandings when information about patients is exchanged and when groups of patients treated by alternative methods are compared or reported from different centres.”

Despite the venerable nature of this fundamental examination technique, studies have shown that there may be limited interrater reliability for estimations of pupil size, shape, and reactivity scores between two clinicians ^16 17^, therefore quantitative pupillometry (QP) has become increasingly popular in the care of a wide variety of patients, particularly in neurocritical care ^18^. QP generally comprises an infrared-sensitive imaging sensor coupled with a digital interface for the automated recording, processing and reporting of pupil data. QP has been shown to allow the detection of subtle early changes in both pupil size and pupillary light reflexes ^19^, and to therefore be reliable in serial quantification of pupil function ^20 21^, particularly following traumatic brain injury with potential cerebral herniation ^22 23^, and in prognostication following cardiac arrest ^24 25 26 27^.

QP facilitates the measurement of 4 phases within the PLR - response latency, maximum constriction, pupil escape and recovery ^28^. Response latency describes the delay in pupil constriction following the beginning of a light stimulus, with increasing light intensity shortening the latency period to a minimum of 180–230 ms, due to both delay in iris smooth muscle contraction and the temporal dynamics of retinal output and innervation pathways ^29 30^. This latency period is followed by a period of rapid constriction of the pupil until it reaches maximum constriction velocity (MCV), after which constriction slows until the minimum pupil diameter is reached ^31^. The maximum constriction amplitude (MCA) is the difference between the baseline and minimum pupil diameter, a computed normalised quantity as a smaller MCA is observed with a smaller baseline pupil diameter ^32^. Following the maximum constriction, the pupil “escapes” to a partially constricted state during a prolonged light stimulus, subsequently undergoing redilation to the initial size in anywhere from 1 to 100 seconds ^33^.

The NPi-200™ uses infrared pupillometry through a light-shielding eye cover on a single eye, exposing it to a controlled 0.8-second light stimulus of a specific intensity, then digitally recording pupil size, reaction latency, and the velocity and amplitude of dilatation and constriction responses at high frequency over 3 seconds post stimulus. The NPi-200™ generates the Neurological Pupillary index (NPi), which incorporates PLR measurements into a single value using a proprietary formula, within the scale of 0-5, with 3-5 being considered “normal”, <3 abnormal, and 0 a fixed, non-reactive pupil. This is said to index the responses to initial pupil diameter, despite the effect of confounders like ambient light or opiate administration.

The MindMirror™ smartphone application is a novel automated pupillometer that adapts a standard smartphone camera for portable user-friendly PLR assessment. Using the native camera, the application records a video of the PLR in response to flash stimulus, recorded over 6 seconds. The application does not require a light shield, delivers binocular imaging, and provides feedback to the user about the ideal distance to hold the camera from the subject to optimise measurement quality.

## Correlation

There were observable differences in minimum / maximum values between the NPi 200™ and MindMirror™ curves, which are potentially explained by the fact that NPi 200™ uses a much brighter flash, potentially triggering a more dramatic initial pupil response., Initial and final pupil diameters, average and maximum constriction velocity, and latency, have all been demonstrated to decrease with increasing light intensity, whereas average and maximum dilation velocities, constriction ration and time taken for 75% recovery to the initial pupil diameter (T75) have all been shown to increase ^34^.

Pearson correlation coefficients between NPi 200™ and MindMirror™ demonstrated a high degree of correlation between both devices for the same measured variables, with narrow standard deviations, and including maximum and average constriction velocities to a light stimulus, and maximum and average dilation velocities following light-induced constriction. This suggests that the MindMirror™ smartphone application technology has similar precision to the NPi 200™, which is a TGA-approved clinical device in Australia, and which is also 510k exempt by the US FDA and CE-marked for the European Economic Area.

### Agreement

Correlation focuses on the association of changes in two outcomes, however, with a high degree of correlation suggesting that the variance of the measured variables are strongly related to each other, reflecting the strength of the association between them. To ensure that the NPi 200™ and MindMirror™ application are providing a similarly accurate measurement of PLR component variables, however, we utilised Bland-Altman agreement ^35^.

Bland–Altman agreement plots are used widely in method comparison studies with quantitative outcomes, where pairs of observations are recorded from the same subject using two different methods, then both means and differences of these pairs of values being displayed in a scatter-plot showing limits of agreement ^36^. The mean difference is displayed, and the limits of agreement represent boundaries within which approximately 95% of all population differences lie.

In this study, agreement between both methods was very good, with a small persistent bias towards a lower absolute measurement of CV, MCV and DV using the MindMirror™ application, compared with the NPi 200™. CV measured in the L eye showed a mean difference of 0 mm s^-1^, and in the R eye a difference of approximately 0.3 mm s^-1^, with the spread of difference values within the 1.96 SD limits of agreement. A difference of approximately 4 mm s^-1^ was seen in MCV, once again with the spread of difference values mostly well within the 1.96 SD limits of agreement. DV measured in both eyes showed a very narrow spread of differences mostly clustered around small mean differences of 0.3 mm s^-1^ in the L eye and 0.1 mm s^-1^ in the R. Although these differences are small, systematic disparities in velocities between L and R eyes are physiologically implausible among a number of unrelated volunteers, therefore it is likely that these differences are artifactual and related both to small study numbers. Later studies are planned with larger numbers to mitigate some potential sources of error.

From the experimental design point of view, it is also important to note that raw data from the NPi 200™ was unavailable, and therefore true precision was challenging to estimate. Investigators chose to prepare a NPi 200™ dataset by manually tracing key points over the screen capture of the device’s graphical output of pupil curves, using a very high resolution camera, however this may in itself have introduced a potential non-systematic source of error.

Despite the apparent tendency of the MindMirror™ to measure the maximum constriction and dilation velocities at a higher value than the NPi 200™, it is important to remember that a systematic bias that is present and similar in all measurements may be an acceptable situation, as reproducible changes in velocities in proportion to the presence and degree of pathology are the fundamental findings that are essential to identify and quantify. A non-systematic error would be far more problematic as this would mean that the direction of bias would be unpredictable and could therefore invalidate the approach.

In fact, since the objective in the observation and quantification of the components of the pupillary light reflex is to precisely identify changes that may signify underlying pathology, correlation between the two devices is critically important. Correlation describes the strength of the relationship between two measurements, and therefore an excellent value for correlation at around 0.9 demonstrates that covariance between these measures is robust and stable.

The ability to identify changes in component PLR velocities, as well as latency, initial and final pupil diameters, and other potential metrics recognised to represent head-injury and concussion-related effects, requires reproducible and robust measurement of highly variable physiological responses. The MindMirror™ smartphone application has been shown to deliver variables highly correlated with existing measurement tools. As a potentially ubiquitous smartphone application, MindMirror™ can be available to all grades of athlete and their responsible staff, whether amateur or professional, and across ages, demographies and locations; and can be used in a plethora of community settings, even in first aid, educational and family care situations.

Further validation and large-scale population-based studies are under way, which will demonstrate MindMirror™’s capacity to deliver almost instantaneous diagnostic and prognostic insights into situations replete with potentially disastrous possibilities.

## Data Availability

All data produced in the present study are available upon reasonable request to the authors

## Statements and Declarations

- Non-financial interests: Professor Middleton and Dr Davies act as unpaid medical advisors to MindMirror inc.
- Financial interests: Mr. Buzytsky is a cofounder and CTO of MindMirror, Inc.

## Ethics approval

This study was performed in line with the principles of the Declaration of Helsinki, and following approval from the South Western Sydney Local Health District (SWSLHD) Human Research Ethics Application (HREA), under the title “2023/ETH00741: The Australian Pupillary Light Response Registry (APLRR)”. Informed consent was obtained from all volunteers.

